# The COVID-19 pandemic and ophthalmic care: a qualitative study of patients with neovascular age-related macular degeneration (nAMD)

**DOI:** 10.1101/2021.09.01.21262696

**Authors:** Seán R O’Connor, Charlene Treanor, Elizabeth Ward, Robin A Wickens, Abby O’Connell, Lucy A Culliford, Chris A Rogers, Eleanor A Gidman, Tunde Peto, Paul C Knox, Benjamin J L Burton, Andrew J Lotery, Sobha Sivaprasad, Barnaby C Reeves, Ruth E Hogg, Michael Donnelly, MONARCH Study Group

**Affiliations:** Centre for Public Health, Queen’s University of Belfast, Belfast, BT12 6BA, UK; Bristol Trials Centre (CTEU), University of Bristol, Bristol Royal Infirmary, Bristol, BS2 8HW, UK; Exeter Clinical Trials Unit (EXECTU), University of Exeter, St Lukes Campus, Exeter, EX1 2LT; Department of Eye and Vision Science, University of Liverpool, Liverpool, L7 8TX, UK; James Paget University Hospitals NHS Foundation Trust, Norfolk, NR31 6LA, UK; Department of Clinical and Experimental Sciences, Faculty of Medicine, University of Southampton, Southampton, SO16 6YD, UK; NIHR Moorfields Biomedical Research Centre, Moorfields Eye Hospital NHS Foundation Trust, London, EC1V 2PD, UK

## Abstract

**Background/aims:** Concerns have been expressed about the relationship between reduced levels of health care utilisation and the COVID-19 pandemic. This study aimed to elicit and explore the views of patients with neovascular age-related macular degeneration (nAMD) regarding the COVID-19 pandemic and their ophthalmic care.

**Methods:** Between April 29th and September 4th 2020, semi-structured telephone interviews were conducted with thirty-five patients with nAMD taking part in a larger diagnostic accuracy study of home-monitoring tests. Participants were recruited using maximum variation sampling to capture a range of key characteristics including age, gender and time since initial treatment. Transcribed interview data were analysed using a deductive and inductive thematic approach.

**Results:** Three themes emerged from the analysis: i. access to eye clinic care. ii. COVID-19 mitigating factors and care delivery and iii. social and personal circumstances. Participants reported anxieties about cancelled or delayed appointments, limited communication from clinic-based services about appointments, and the impact of this on their ongoing care. Despite these concerns, there was apprehension about attending appointments due to infection risk and a perception that nAMD patients are a ‘high risk’ group. Views of those who attended clinics during the study period were, however, positive, with social distancing and infection control measures providing reassurance.

**Conclusions:** These findings contribute to our understanding about experiences of patients with nAMD during the COVID-19 pandemic and have potential implications for future planning of care services. Innovative approaches may be required to address issues related to access to care, including concerns about delayed or cancelled appointments.

**Synopsis:** Perspectives of patients with neovascular age-related macular degeneration regarding the COVID-19 pandemic identified important issues regarding access to, and experience of ophthalmic care. These findings have implications for future planning of services.

## BACKGROUND

The World Health Organization (WHO) formally declared Coronavirus disease 2019 (COVID-19) as a pandemic on 11 March 2020.^1^ COVID-19 is an infectious acute respiratory disease caused by a novel coronavirus (SARS-CoV-2).^2^ Older people and those with underlying health conditions are at increased risk of developing more serious illness that may have significant longer-term health effects.^3^ Ophthalmology clinics may be impacted by COVID-19, as clinicians often perform assessments, examinations and deliver treatments to patients in close proximity. Methods of remote care in place of clinic-delivered treatment may therefore be less viable in comparison to other areas of clinical practice.^4^ As a result of this and other structural factors, many clinic services stopped or substantially reduced the usual schedule of clinical appointments and procedures. This decrease in service provision subsequently led to indirect health effects during the COVID-19 pandemic.^5^ Indirect effects tended to be associated with increased patient morbidity because of restricted preventative care services, diagnostic delays and reduced intervention delivery.^6^ More broadly, national level and localised lockdowns, and social distancing measures, contributed further in terms of restricted patient access to ongoing care. Furthermore, public information and advice that was designed to mitigate the effects of COVID-19 transmission appeared to influence a perception that attending a hospital appointment or an unscheduled clinic visit could increase the risk of contracting the virus.^7^ Neovascular age-related macular degeneration (nAMD) is a chronic, progressive condition and the commonest cause of vision loss in older adults,^8^ with global prevalence predicted to increase from 196 million in 2020, to 288 million in 2040.^9^ Ongoing surveillance is necessary to manage disease activity since nAMD can recur following periods of treatment.^10^ Therefore, it is important to examine any changes to clinic-based services in relation to patient outcomes. This qualitative study elicited and explored the views of patients with nAMD regarding the COVID-19 pandemic and changes to their ophthalmic care.

## MATERIALS AND METHODS

Qualitative methods were used to explore patients’ responses, views and experiences, and to examine variations in personal contexts.^11^ Participants were interviewed at least one month after the COVID-19 pandemic was declared by the WHO^1^ and once public health measures were in place in the UK.^12^ This was during the first UK he study followed the consolidated criteria for reporting qualitative research (COREQ).^13^ Ethical approval was acquired from the National Research Ethics Service (IRAS ref: 232253 REC ref: 17/NI/0235).

### Participants

Remote, semi-structured interviews were conducted between April 29th and September 14th 2020, covering the period during the first national lockdown in March 2020 and the easing of restrictions in August 2020. The study included a subset of nAMD patients who were taking part in a larger diagnostic accuracy study of home-monitoring tests (MONARCH).^14^ Participants were recruited from four sites within the UK. Maximum variation sampling was used to ensure that a range of perspectives were captured in relation to age category (young-old 50-69 years and older-old 70+years), gender, laterality of nAMD (unilateral and bilateral) and time since first treatment (6-17 months, 18-29 months and 30-41 months). Participants were given a minimum of one week to consider the study information and discuss it with family members before agreeing to take part. Informed consent to participate in the study was obtained verbally prior to interviews and following a full explanation of study procedures.

### Data collection

All participants were given the option of completing the semi-structured interview using video-conferencing software, or via telephone. The interview schedule (see Supplementary file 1) was developed based on the experience of the research team and and was informed by relavent theoretical models including the Theoretical Framework of Acceptability.^15^ Members of the team who collected and analysed the data (CT, SOC, MD) had extensive experience in the application of qualitative methods in healthcare research. No participant was known to the researchers who conducted the interviews.

### Data analysis

Interviews were audio-recorded and transcribed. A directed content analysis approach based on deductive and inductive coding was used.^16^ Coding underwent iterative development as individual transcripts were reviewed and re-reviewed during data familiarisation (CT, SOC, MD) (See Supplementary file 2). Following line-by-line coding of each transcript (CT, SOC), findings related to views on usual care, the impact of COVID-19 on care, and views about the COVID-19 pandemic in general were summarised. Transcripts were cross-coded and discussed to ensure rigour and reflexivity. Related codes were clustered and grouped into themes that were reviewed and refined to ensure coherence. NVivo version 12 was used to manage data and facilitate the analysis process. This process, in summary, included the following stages: i. independent transcription, ii. Data familiarisation, iii. independent coding, iv. development of an analytical framework, v. indexing, vi, charting and vii. interpreting data.

## RESULTS

Two participants who were approached declined to take part. A total of 35 interviews were completed. Interviews took place in the context of organisational and structural changes to clinic services including the cancelation of routine appointments and prioritisation of urgent care. In some cases, virtual methods were used to ‘triage’ patients and identify if there was a need to attend an urgent appointment. All participants opted to complete the remote interviews by telephone. The demographics of participants are shown in Table 1. The majority of participants were female (69%) with a mean age of 77 years. Interviews lasted an average of 48 minutes (range: 39 to 78 minutes).

**Table 1.**
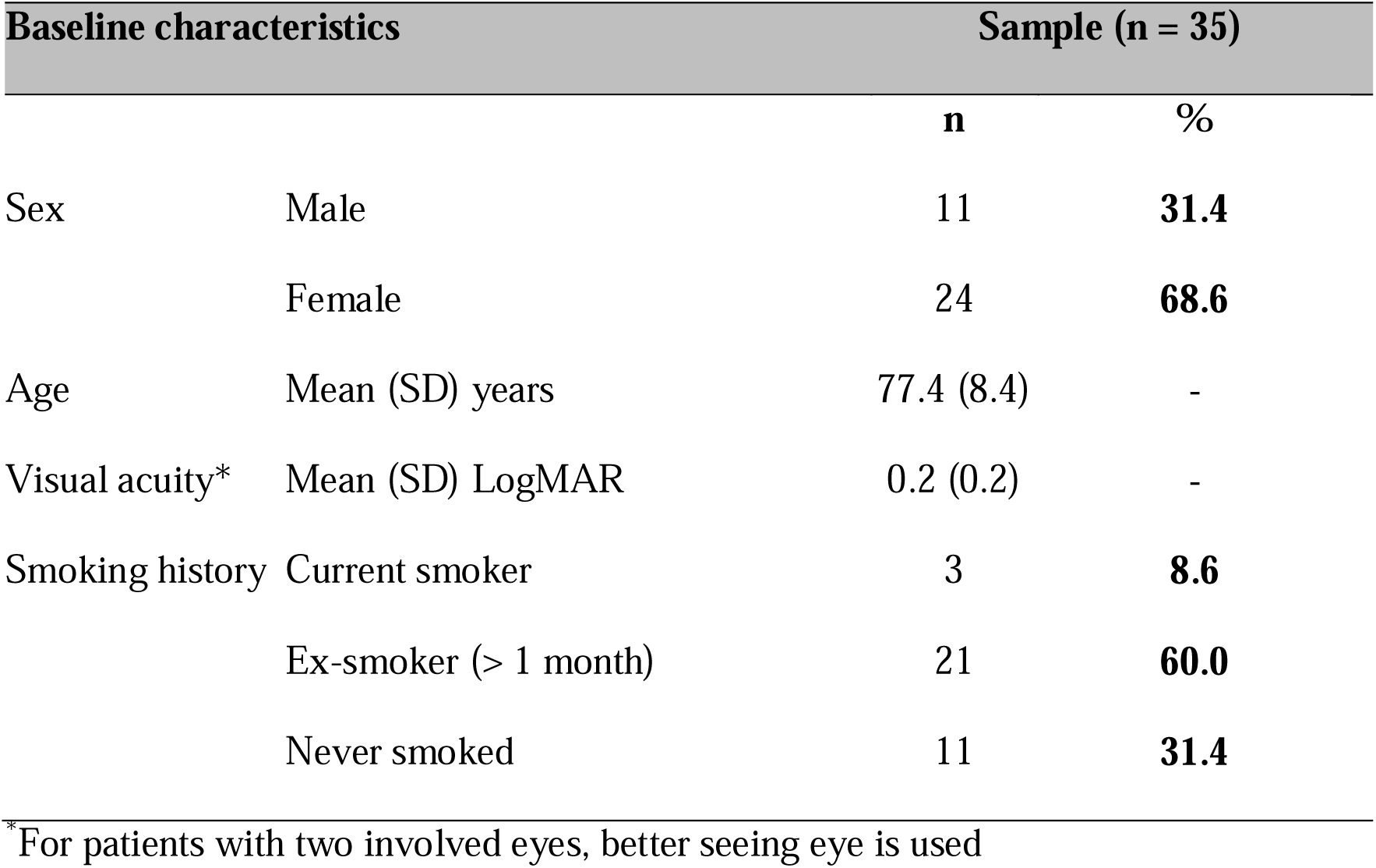
Demographic characteristics of participants.

| Baseline characteristics |  | Sample (n = 35) |  |
| --- | --- | --- | --- |
|  |  | n | % |
| Sex | Male | 11 | **31.4** |
|  | Female | 24 | **68.6** |
| Age | Mean (SD) years | 77.4 (8.4) | - |
| Visual acuity* | Mean (SD) LogMAR | 0.2 (0.2) | - |
| Smoking history | Current smoker | 3 | **8.6** |
|  | Ex-smoker (> 1 month) | 21 | **60.0** |
|  | Never smoked | 11 | **31.4** |

\*For patients with two involved eyes, better seeing eye is used

With respect to their experiences of COVID-19, one participant reported having had a negative test for COVID-19 and two reported that they had experienced suspected coronavirus related symptoms previously. Three participants were advised to self-isolate due to close contact with a person diagnosed with COVID-19. No participants were diagnosed with COVID-19 at the time of their interview.

Three overarching themes emerged from the analysis and revolved around: i. access to eye clinic care, ii. COVID-19 mitigating factors and care delivery, and iii. social and personal circumstances. Aspects of the three themes overlapped. Each theme is presented in the section below. Selected illustrative quotes from participants are shown in Tables 2 and 3.

**Table 2.**
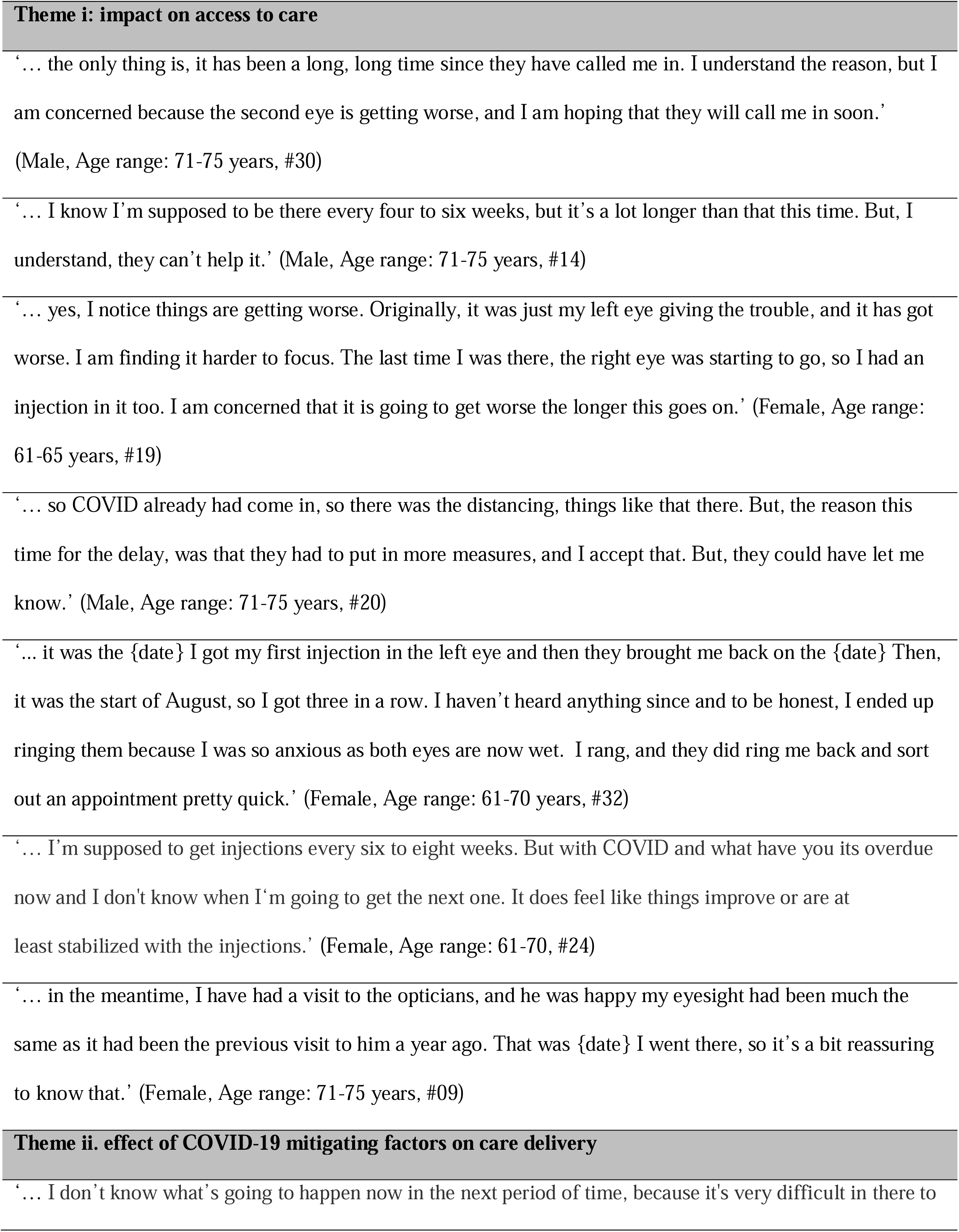
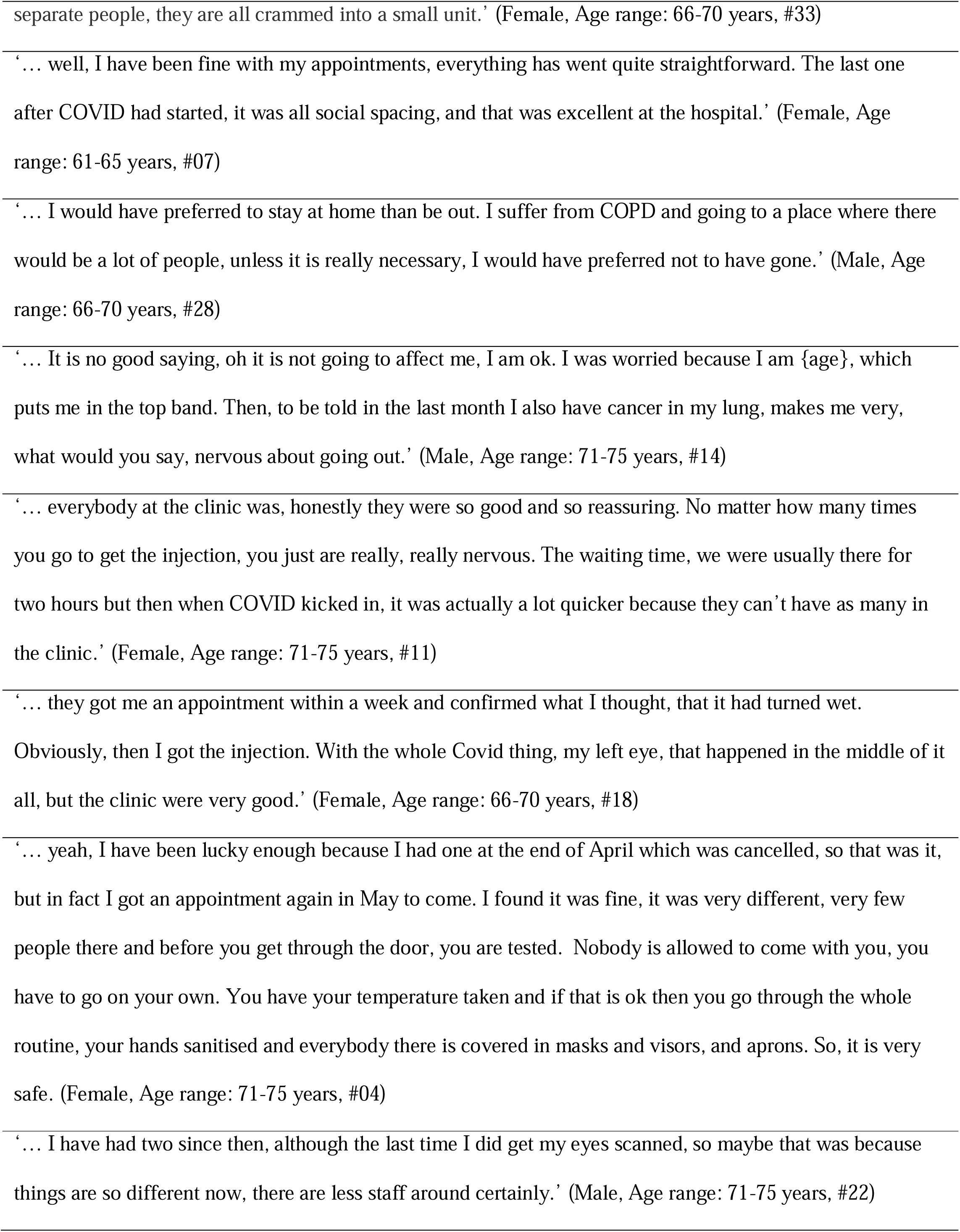
Selected quotes related to views on the impact of COVID-19 on access to care and the effect of mitigating factors on care delivery.

**Table 3.**
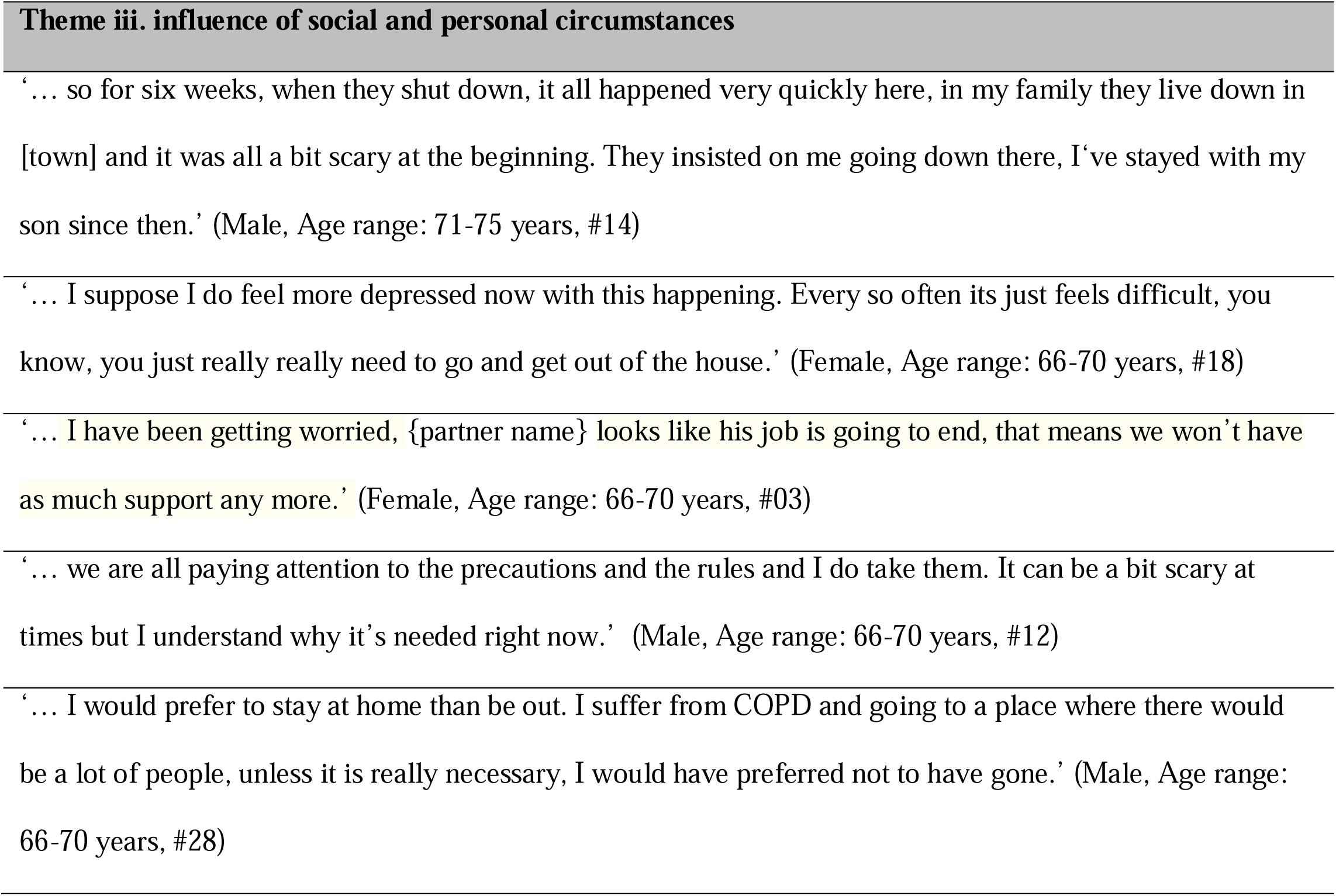
Selected quotes related to views on the influence of COVID-19 on social and personal circumstances.

### Theme i. access to eye clinic care

Participants became concerned when their access to eye clinic care was restricted as a result of changes to services during the COVID-19 pandemic and worried that clinics would remain closed during the pandemic – the vast majority had expected or previously arranged appointments cancelled by the time of their qualitative research interview. Greater concerns about not attending an appointment were apparent among patients who described having attended clinic appointments at regular intervals (e.g. every four to six weeks) before the pandemic and who were, therefore, expecting to attend an appointment when changes to clinical services were implemented. More specifically, patients reported fears about potential worsening of symptoms or deterioration in their vision and uncertainty about what to do in this event. Some participants described how they felt that deterioration had occurred. This was reported typically by patients who had continued to receive treatment during ther pandemic, and was therefore attributed by these patients to normal disease progression and not to the impact of the pandemic on services. It was also highlighted that participants missed the face-to-face contact and reassurance that clinic visits provided, but the need for organisational changes due to infection risk during the the pandemic, resulting in limited access to services, was acknowledged. It was also assumed that these changes were a result of clinical staff being temporarily redirected to other areas, as this was something that participants described noticing in other areas of the health service.

The importance and value of effective communication was emphasised particularly in relation to information for patients about delayed appointments and their rescheduling. Actual experiences of communication from clinics were mixed. Some participants, who had not been contacted about appointments, were unsure why this was the case, and were not aware that prioritisation of urgent care was in place. In some cases, participants had initiated contact with services and enquired about the rescheduling of appointments, though others did not feel able or willing to contact their clinic. Those participants who contacted services tended to be younger and female.

Although no participants had taken part in any remotely delivered or virtual eye clinic appointments, some had done so in other care contexts, primarily as part of primary care. It was highlighted that these methods could be a possible, albeit temporary solution which might be used by services to enquire about and ‘assess’ any changes in vision. Other ‘models’ of care were suggested by participants to ensure access to ongoing care, included the use of community-based optometry services for assessing nAMD progression. These treatment delivery arrangements were also viewed as possible ways in which to relieve the burden on hospital eye services beyond the COVID-19 pandemic.

### Theme ii. COVID-19 mitigating factors and care delivery

While there were fears around deterioration in vision, or worsening of other symptoms, participants were often apprehensive about attending hospital appointments as part of their eye care, or for management of co-morbidities or other existing conditions. This was due to the perceived risk of COVID-19 infection, and the risk of severe illness if infected due to the older age of nAMD patients. Despite this, the opinions of the small number of patients (approximately one third) who did attend eye clinic appointments during the study period were typically very positive. Participants highlighted the professionalism of clinic staff, and felt reassured that mitigating factors including social distancing and infection control measures were being strictly adhered to. Appointments were compared to the experience of visiting clinics prior to the pandemic. Previous visits were often described as being lengthy, involving long waiting times and held in busy and sometimes crowded environments. As a result of COVID-19 mitigating factors, appointments during the pandemic were seen as involving markedly shorter waiting times, clear social distancing and with appropriate use of personal protective equipment. One negative aspect for participants was that partners or relatives who usually attended appointments with patients were unable to do so and therefore could not provide support during visits, or if usually provided, practical assistance such as providing transport to clinic appointments. An additional concern raised was that participants worried about the risk of partners or family members contracting coronavirus infection after they had visited hospital sites.

### Theme iii. social and personal circumstances

Participants reported experiencing a sense of social isolation, as well as generalised anxiety related to the impact of the pandemic, and ambiguity about the long-term effects on their care and overall quality of life. Many described how the pandemic had a significant effect on their routines and felt that lockdowns and social distancing requirements, and the need to self-isolate, had a major influence on their social interactions and in some cases, employment status. In a few cases, participants described having to move temporarily and live with other family members. It was felt by some that measures were applied without sufficient information being provided around how long they might be in place, and for these participants, this increased their sense of uncertainty. Despite feeling that health information was conflicting, participants also felt strongly about adhering to health measures. This was most apparent in those participants with comorbidities, including cancers, and respiratory conditions who stated that they felt at increased risk. There was a clear understanding around the reasons for health measures, even if it resulted in changes to routines, and views were strongly negative towards those who do not adhere to public health measures.

## DISCUSSION

In the present study, initial evidence is provided which can contribute towards an improved understanding of the impact of the COVID-19 pandemic on patients with nAMD. The importance of conducting qualitative research during the COVID-19 pandemic has been highlighted.^17^ Qualitative methods are essential to provide in-depth exploration of patient perspectives and can also be used alongside longitudinal or retrospective study designs to assess the effect of COVID-19 on patient outcomes.

Three themes emerged from the analysis. These related to access to care, the effect of mitigating factors on care delivery, and the influence of patients’ social and personal circumstances. Concerns were reported about limited access to care, and missed or delayed eye clinic appointments, but there was a common understanding around the reasons for the organisational changes to services because of the pandemic. Participants reported experiencing a sense of social isolation, as well as generalised anxiety related to the impact of the pandemic, and uncertainty about the long-term effects on their care and overall quality of life. While there were fears around deterioration in vision, or worsening of other symptoms, participants were also often apprehensive about attending hospital appointments as part of their eye care, or for management of co-morbidities or other existing conditions. This apprehension was related to perceived infection risks and a view that nAMD patients as an older population were therefore a ‘higher risk’ group. An additional concern raised was that participants worried about the risk to partners or family members of contracting coronavirus infection after they had visited hospital sites. Despite these observations, the opinions of patients who did attend clinical appointments during the study period were positive. Participants highlighted the professionalism of clinic staff, and their strict adherence to measures to mitigate the risk of infection, including social distancing and personal protective equipment. In an earlier study,^18^ information and support, as well as additional factors which influence service delivery, such as appointment and waiting times, were highlighted as potential targets to improve patients’ experience of being assessed for and receiving treatment for AMD. In the present study, the importance and value of effective communication with services was also emphasised. This was seen as important particularly in relation to patients being provided with information on delayed appointments, and when they might be rescheduled. It was interesting that only some participants felt it was important to contact services to enquire about future appointments and others did not. This may have been related to the stage of nAMD, e.g. whether patients were receiving active treatment before the pandemic or surveillance only. These different reactions to their circumstances would merit more detailed examination.

Our findings are broadly reflective of those reported in other studies examining the impact of COVID-19 in patient populations. Concerns about restricted access to care and social isolation as a result of the pandemic have been reported in different groups, including those with chronic pain disorders,^19^ diabetes^20^ and obesity^21^. Other studies have confirmed that the pandemic has been associated with reductions in routine assessments and treatment for various conditions,^22^ as well as in ophthalmology care contexts.^5,6^ Going forward, the need for reorganization of services to reduce the effects of these changes to services on patient outcomes is acknowledged.^23^ For example, it is recognised that it is likely that the need for remotely delivered care will continue to increase.^24^ Other models of care may also be required, including increased use of community-based optometry services for managing nAMD, to relieve burden on hospital eye service. However, potential barriers to use of such services have been previously identified, including concerns about potential delays in referrals for intervention when it is required.^25^

A potential limitation of the study is that use of remote telephone interviews may provide different information than would be gathered using face-to-face interviews.^26,27^ This method was however, precluded because of the COVID-19 pandemic and associated social distancing measures. The study was also based on interviews conducted with participants taking part in an ongoing diagnostic accuracy study. Questions around the impact of COVID-19 were therefore not the only focus of the interviews and the responses may have been less in depth than if they related only to COVID-19. While participants were recruited to the study using maximum variation sampling methods (to ensure a balance of important patient characteristics), the sample may also not be reflective of all patients with nAMD. Participants were also recruited from sites within the UK and findings may not be applicable to other healthcare systems. Another limitation is that interviews could have been influenced by the different phases of the pandemic at which the interviews were conducted.

To our knowledge, this is the first study to report on the perspectives of nAMD patients regarding the impact of the COVID-19 pandemic on their care. Three themes emerged from the analysis related to concerns about access to care, the effect of mitigating factors on care delivery, and the influence of patients’ individual circumstances. The most significant factor was the impact on access to care. Participants emphasised the importance and value of effective communication by services to address these concerns. Participants also highlighted how alternative models of care could play a part in managing issues around access to care. This included remote methods of delivery and increased use of other models of care such as optometry services to reduce the burden on ophthalmology services at times of strain on hospital based systems. In summary, these findings could be used to understand the experiences of patients with nAMD during the ongoing COVID-19 pandemic, and could have implications for future planning of care services in the event of subsequent waves, or future pandemics. Innovative approaches may be required to address the issues raised related to patients’ concerns about ensuring adequate access to care. Consideration should also be given to supporting patients to manage social isolation and anxiety in vulnerable patient groups, including those in older populations, and those who have existing co-morbidities or chronic health-related conditions such as nAMD. Further studies examining the indirect health effects of the COVID-19 pandemic in ophthalmology are also essential to improve understanding of its impact on longer-term patient and service level outcomes.

## Data Availability

n/a

## Acknowledgements

The study is sponsored by The Queen’s University of Belfast, UK. This study was designed and delivered in collaboration with the Clinical Trials and Evaluation Unit (CTEU), a UKCRC registered clinical trials unit which, as part of the Bristol Trials Centre, is in receipt of National Institute for Health Research CTU support funding. The authors would like to acknowledge the Patient and Public Involvement group (PPI), the Macular Society and the Royal National Institute of Blind People for feedback on the study and the patient documents and the SSC for their oversight of the study.

## Authors’ contributions

REH and BCR developed the original proposal for the diagnostic accuracy study and were co-chief investigators; MD developed the qualitative study component of the diagnostic study. REH, BCR, MD, AJL, SS, PCK, BJLB and CAR obtained funding. MD, SOC and CT were responsible for data collection, analysis and interpretation. EW, RAW, AOC, LAC, CAR, EAG provided expert input. TP, AJL, SS, PCK and BJLB provided clinical expertise and input. SOC and MD drafted the initial manuscript and all authors revised the manuscript for important intellectual content and approved the final version.

## Funding

This project was funded by the National Institute for Health Research, Health Technology Assessment (HTA) Programme (ref 15/97/02). The views and opinions are the authors and do not necessarily reflect the HTA programme, NIHR, NHS or the Department of Health and Social Care.

## Conflict of interest

The authors declare that they have no competing interests.

## Patient consent

Informed consent was obtained in accordance with the arrangements that were approved by the Ethics Committee (see below).

## Ethics approval

Ethical approval was acquired from the National Research Ethics Service (IRAS ref: 232253 REC ref: 17/NI/0235).

---

**Supplementary file 1.**
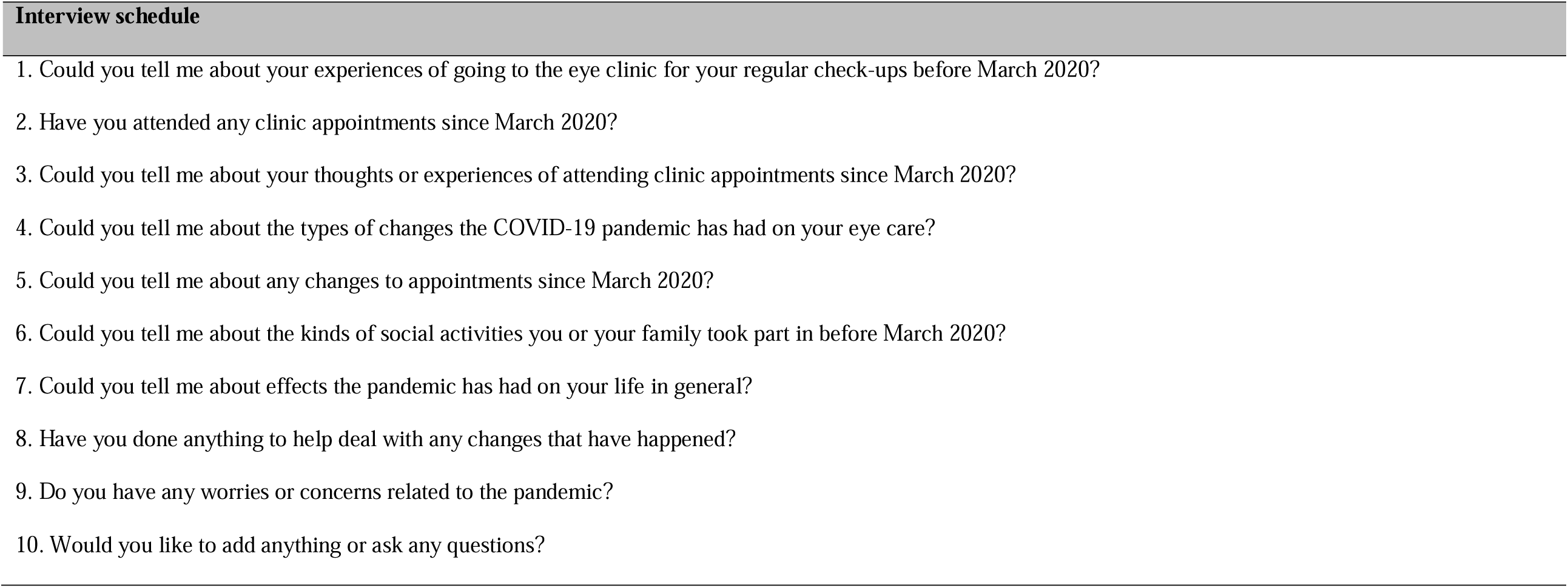
Interview schedule.

**Supplementary file 2.**
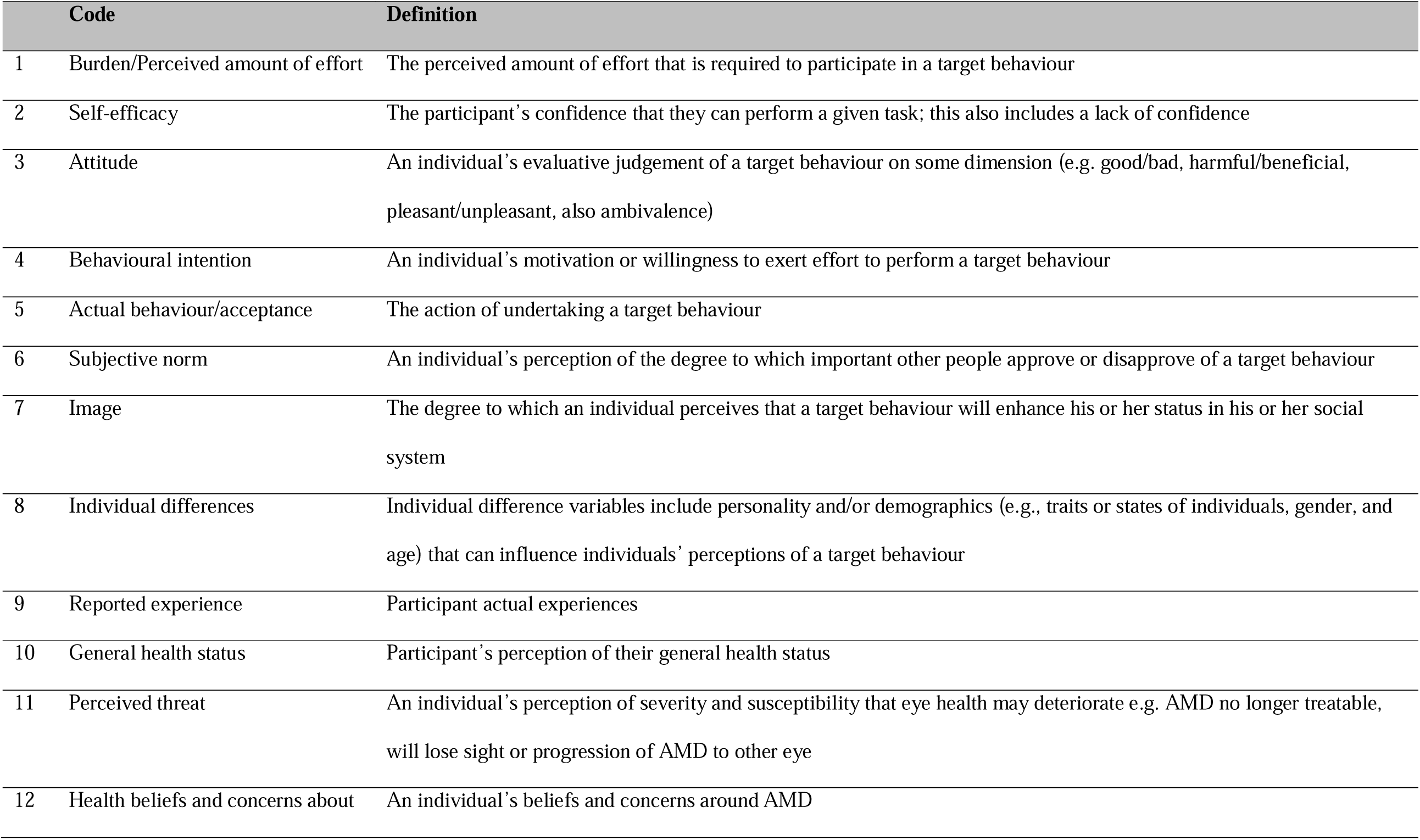
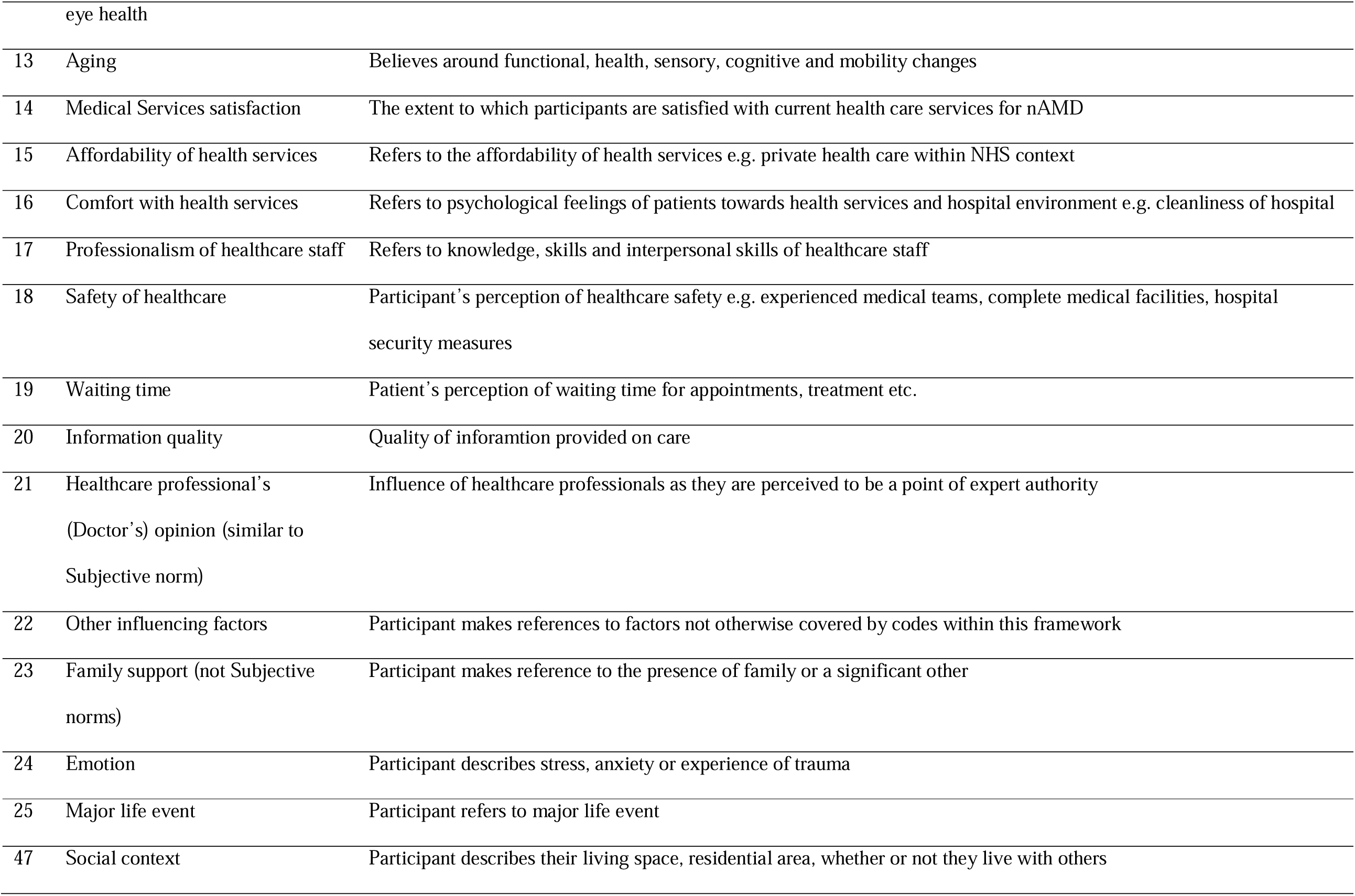
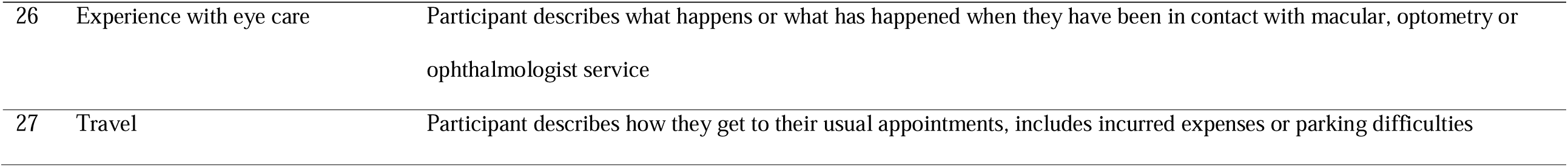
Coding framework.

## Notes

### Competing Interest Statement

The authors have declared no competing interest.

### Clinical Trial

n/a

### Author Declarations

Ethics approval Ethical approval was acquired from the National Research Ethics Service (IRAS ref: 232253 REC ref: 17/NI/0235).

