## Supplementary file 1 for "The COVID-19 pandemic and ophthalmic care: a qualitative study of patients with neovascular age-related macular degeneration (nAMD)"

**Supplementary file 1. Interview schedule**

**Interview schedule**

1. Could you tell me about your experiences of going to the eye clinic for your regular check-ups before March 2020?

2. Have you attended any clinic appointments since March 2020?

3. Could you tell me about your thoughts or experiences of attending clinic appointments since March 2020?

4. Could you tell me about the types of changes the COVID-19 pandemic has had on your eye care?

5. Could you tell me about any changes to appointments since March 2020?

6. Could you tell me about the kinds of social activities you or your family took part in before March 2020?

7. Could you tell me about effects the pandemic has had on your life in general?

8. Have you done anything to help deal with any changes that have happened?

9. Do you have any worries or concerns related to the pandemic?

10. Would you like to add anything or ask any questions?
