## Supplementary file 2 for "The COVID-19 pandemic and ophthalmic care: a qualitative study of patients with neovascular age-related macular degeneration (nAMD)"

**Supplementary file 2. Coding framework**

|  | **Code** | **Definition** |
| --- | --- | --- |
| 1 | Burden/Perceived amount of effort | The perceived amount of effort that is required to participate in a target behaviour |
| 2 | Self-efficacy | The participant’s confidence that they can perform a given task; this also includes a lack of confidence |
| 3 | Attitude | An individual’s evaluative judgement of a target behaviour on some dimension (e.g. good/bad, harmful/beneficial, pleasant/unpleasant, also ambivalence) |
| 4 | Behavioural intention | An individual’s motivation or willingness to exert effort to perform a target behaviour |
| 5 | Actual behaviour/acceptance | The action of undertaking a target behaviour |
| 6 | Subjective norm | An individual’s perception of the degree to which important other people approve or disapprove of a target behaviour |
| 7 | Image | The degree to which an individual perceives that a target behaviour will enhance his or her status in his or her social system |
| 8 | Individual differences | Individual difference variables include personality and/or demographics (e.g., traits or states of individuals, gender, and age) that can influence individuals’ perceptions of a target behaviour |
| 9 | Reported experience | Participant actual experiences |
| 10 | General health status | Participant’s perception of their general health status |
| 11 | Perceived threat | An individual’s perception of severity and susceptibility that eye health may deteriorate e.g. AMD no longer treatable, will lose sight or progression of AMD to other eye |
| 12 | Health beliefs and concerns about eye health | An individual’s beliefs and concerns around AMD |
| 13 | Aging | Believes around functional, health, sensory, cognitive and mobility changes |
| 14 | Medical Services satisfaction | The extent to which participants are satisfied with current health care services for nAMD |
| 15 | Affordability of health services | Refers to the affordability of health services e.g. private health care within NHS context |
| 16 | Comfort with health services | Refers to psychological feelings of patients towards health services and hospital environment e.g. cleanliness of hospital |
| 17 | Professionalism of healthcare staff | Refers to knowledge, skills and interpersonal skills of healthcare staff |
| 18 | Safety of healthcare | Participant’s perception of healthcare safety e.g. experienced medical teams, complete medical facilities, hospital security measures |
| 19 | Waiting time | Patient’s perception of waiting time for appointments, treatment etc. |
| 20 | Information quality | Quality of inforamtion provided on care |
| 21 | Healthcare professional’s (Doctor’s) opinion (similar to Subjective norm) | Influence of healthcare professionals as they are perceived to be a point of expert authority |
| 22 | Other influencing factors | Participant makes references to factors not otherwise covered by codes within this framework |
| 23 | Family support (not Subjective norms) | Participant makes reference to the presence of family or a significant other |
| 24 | Emotion | Participant describes stress, anxiety or experience of trauma |
| 25 | Major life event | Participant refers to major life event |
| 47 | Social context | Participant describes their living space, residential area, whether or not they live with others |
| 26 | Experience with eye care | Participant describes what happens or what has happened when they have been in contact with macular, optometry or ophthalmologist service |
| 27 | Travel | Participant describes how they get to their usual appointments, includes incurred expenses or parking difficulties |
